# Interpretable Symptom-Based Machine Learning for Parkinson’s Disease Prediction: A Feasibility Study

**DOI:** 10.64898/2026.05.15.26352866

**Authors:** Md Zainul Ali, Pankaj Singh Dholaniya

**Author notes:** **Corresponding Author: Pankaj Singh Dholaniya, Ph.D.** Assistant Professor, Department of Biotechnology and Bioinformatics, School of Life Sciences, University of Hyderabad.

## Abstract

**Background:** Parkinson’s disease (PD) has a prolonged prodromal phase during which non-motor symptoms (NMS) may emerge years before the appearance of classical motor signs. This makes NMS a promising and clinically accessible source of information for early risk stratification.

**Objective:** In this study, we investigated whether NMS alone can serve as reliable predictors of PD risk using clinical data from the Parkinson’s Progression Markers Initiative (PPMI) cohort.

**Method:** We developed a stacked ensemble machine learning framework that integrates feature-level modelling, a global multivariate model, and a patient-similarity component to capture complementary patterns within NMS profiles. The model was trained using leakage-controlled patient-level validation and evaluated on an independent held-out test set.

**Results:** The final ensemble achieved strong predictive performance, with an area under the ROC curve of 0.994, balanced accuracy of 0.9484. Explainability analysis further showed that olfactory dysfunction, gastrointestinal symptoms, urinary and other autonomic features, and selected cognitive measures were among the most influential predictors. These findings support the hypothesis that NMS are not merely associated features of PD, but can function as meaningful predictors of disease risk even without imaging or biomarker inputs. Additionally, the final validated model is implemented into a web-based research prototype to demonstrate real-time translational feasibility.

**Conclusion:** Overall, this study highlights the predictive value of NMS for PD risk assessment and supports their use in research-oriented early screening frameworks.

## 1. Introduction

Parkinson’s disease (PD) is the second most common neurodegenerative disorder worldwide. It affects more than 6 million people, and this number is expected to nearly double by 2040 [1]. The global burden of PD has increased rapidly in the past two decades, making it one of the fastest growing neurological disorders in terms of prevalence, disability, and mortality [2]. In low- and middle-income countries such as India, the burden is higher due to limited specialist care, delayed referrals, and low awareness at the primary care level [3]. PD is still incurable, and current treatments mainly reduce symptoms rather than modify disease progression [4]. A major challenge in PD is the long delay between disease onset and diagnosis [5]. Clinical diagnosis depends on motor symptoms such as bradykinesia, rigidity, and resting tremor. These appear only after 50-70% of dopaminergic neurons are already lost [6, 7]. This results in a long pre-diagnostic period of about 5 to 20 years [8]. During this period, irreversible neuronal damage occurs. Therefore, any disease-modifying therapy must be given before motor symptoms appear to be effective [9].

The prodromal phase of PD includes several non-motor symptoms (NMS). These include REM sleep behaviour disorder, hyposmia, constipation, depression, anxiety, and cognitive decline [10, 11]. These symptoms can appear many years before motor signs. The Braak staging hypothesis explains this early appearance through the spread of pathology from lower to higher brain regions [12]. Clinical tools such as the Movement Disorder Society-Unified Parkinson’s Disease Rating Scale (MDS-UPDRS) help capture these non-motor features [13]. Identifying high-risk individuals using such features before motor onset can be a useful screening approach [14].

Machine learning (ML) has shown strong potential in analysing complex clinical data and supporting diagnosis and risk prediction [15, 16]. In PD, ML models have been applied to imaging, speech, gait, genetics, and clinical data [17, 18]. Algorithms such as Logistic Regression (LR), and support vector machine (SVM) have shown good performance in classification tasks [19]. However, many models use a single method or data type. This limits their ability to generalise across different patient groups and real-world clinical settings [20]. Clinical NMS datasets are often sparse, because different assessments are not performed at every visit or in every participant. This missingness can reduce the stability of multivariate models and motivates approaches that can operate under partial input while maintaining reliable probability estimates. In addition, for screening and risk stratification, probability calibration is important, because outputs are interpreted as risk rather than only as a binary class. Ensemble learning combines multiple models to improve performance and stability [21]. Stacking is a meta-learning approach where outputs from base models are used as inputs for a higher-level model [22]. This helps combine strengths of different algorithms. Collaborative Filtering (CF), commonly used in recommender systems, can also be applied in healthcare. It uses patient similarity to make predictions even when data is incomplete [23]. However, combining these approaches in a single framework for PD risk prediction is still limited. We have utilized the dataset from the Parkinson’s Progression Markers Initiative (PPMI). It is a large longitudinal study that collects detailed clinical, cognitive, imaging, and biological data [24]. Its rich dataset is suitable for developing ML models for PD risk prediction using non-motor features.

In this study, we present *PD-INSPECT* (**P**arkinson’s **D**isease: **I**ntegrated **N**on-motor **S**ymptom-based **P**rediction and **E**valuation using **C**omputational **T**ool), a stacked ensemble framework for PD risk prediction using NMS data. The model combines three components: a feature-level ensemble of LR, SVM, and XGBoost (M1), a global LightGBM model (M2), and a CF module based on patient similarity (M3). A Logistic Regression meta-learner integrates these outputs using a leakage-free stacking approach. The system is also deployed as a web-based tool for easy use by researchers and clinicians. This study proposes a unified framework that combines multiple modelling strategies for PD risk assessment using PPMI data. The main focus of the study is to evaluate whether NMS can serve as meaningful predictors of PD risk using routinely collected clinical data. A web-based interface was developed only as a feasibility study to support future expansion as more data become available. The working architecture of the proposed model, including the three sub-models and the final ensemble prediction process, is illustrated in Figure 1.

**Fig. 1.**
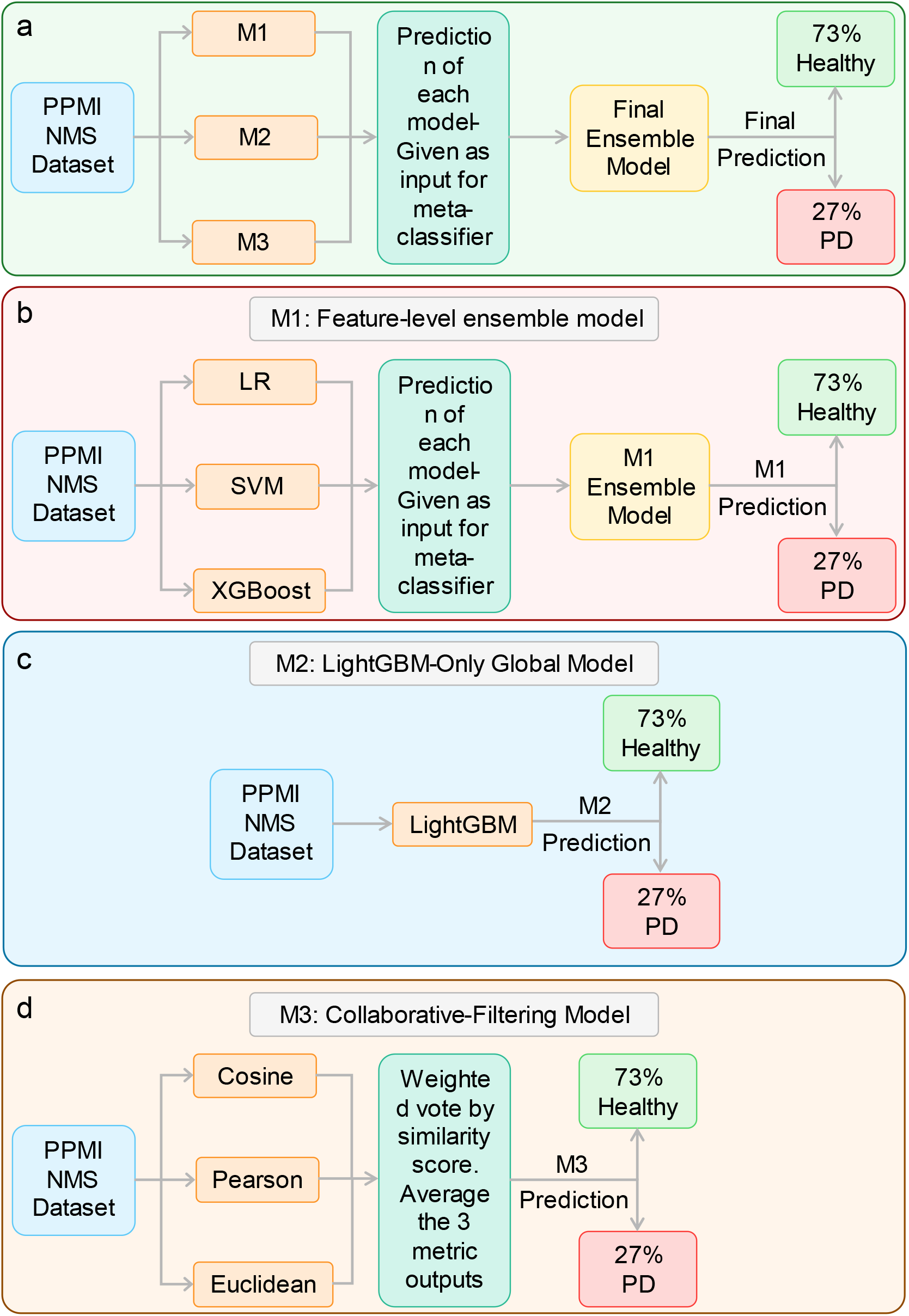
Schematic representation of the modelling framework. (a) Overall study workflow. (b) Architecture of the M1 feature-level ensemble model. (c) Architecture of the M2 global modelling approach using LightGBM. (d) Workflow of the M3 collaborative-filtering model based on patient similarity. The example probabilities shown in the schematic are hypothetical and included solely for visualization of the workflow.

## 2. Methods

### 2.1 Dataset and Pre-processing

Data were obtained from the PPMI, a longitudinal multi-centre observational study designed to identify PD progression markers [24]. The PPMI database provides standardized clinical assessments data collected across multiple follow-up visits from participants of PD and healthy controls populations. NMS represent a clinically relevant feature set in PD because they can appear during the prodromal stage, well before manifestation of motor diagnosis. These features span olfactory, sleep, mood, cognitive, and autonomic domains and can be captured using standardized clinical assessments. Details of the 44 NMS tests used in the present study are presented in Table S1. However, as not all assessments are performed at every visit or in every participant, the resulting NMS dataset is inherently sparse and requires structured pre-processing before modelling. The present work restricted inclusion to participant records with adequate feature coverage, operationalized through two sparsity-filtered data matrices: The first sparse matrix included all sparse data without applying a minimum non-null entry threshold per participant and was used for visit-level modeling in sub-models M1 and M2. The second matrix i.e., 50%-sparse matrix (minimum 50% non-null completeness per participant), used for patient-level modelling in sub-model M3.

Each matrix retained clinical feature scores alongside two demographic covariates, age and sex. Diagnostic labels were encoded as binary values: PD (APPRDX = 1) and healthy control (APPRDX = 2), mapped to 1 and 0, respectively. Sex was re-encoded from the original convention (1 = Male, 2 = Female) to a binary indicator (0 = Male, 1 = Female). For the 50%-sparse matrix, multiple visit records per participant were aggregated to a single patient-level row by computing column-wise means, thereby producing one representative feature profile per patient for use in CF. All 44 clinical features were retained across both matrices after sparsity filtering.

Missing values in the first-sparse matrix were handled at the feature-slice level during model training: training-set medians were computed independently for each (feature, age, sex) slice and applied to both training and validation partitions, ensuring that no information from validation rows influenced imputation parameters. For the 50%-sparse matrix, column-wise training means served as imputation values. Neutral imputation defaults at inference time were computed as the arithmetic mean of the class-specific means (PD mean + Healthy mean) / 2 for each feature, to avoid biasing predictions towards the majority class when a feature is entirely absent in a new patient record. The training data consists of 724 unique participants (565 PD and 159 healthy controls). The held-out test cohort contained 182 patients (142 PD and 40 healthy controls).

### 2.2 Feature space and overall modelling workflow

The input space comprised age, sex, and 44 NMS-centred clinical variables spanning cognition, mood and anxiety, sleep-related symptoms, olfaction, and autonomic domains including gastrointestinal, urinary, cardiovascular, and thermoregulatory measures. All variables were used as numeric inputs without manual feature engineering. The overall workflow was sequential: first, three complementary model families were developed independently; second, multiple variants within each family were benchmarked; third, one final branch was selected from each family; and fourth, the three selected branch-level probabilities were combined in a leakage-controlled logistic-regression meta-learner to generate the final probability of PD [22, 25]. This design was intended to capture complementary predictive structure at the feature-wise, multivariate, and patient-similarity levels.

### 2.3 Development and formulation of the proposed models

#### 2.3.1 Development of M1: feature-level ensemble model

M1 used a two-stage feature-level ensemble strategy. For each visit i and clinical feature j, a three-dimensional input vector z_ij_ = [x_ij_, a_i_, s_i_]^T^ was constructed, where x_ij_ denotes the value of feature j at visit i, a_i_ denotes age, and s_i_ denotes sex. Three classifiers were then trained independently on this feature-specific input: Logistic Regression (LR), a linear Support Vector Classifier (SVC) with isotonic calibration, and XGBoost. The LR branch estimated the probability of PD as:

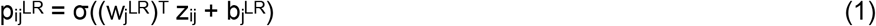

where σ(·) is the sigmoid function. The ranch first generated a decision score, which was then converted to a calibrated probability using isotonic calibration [26]:

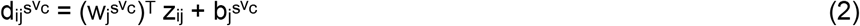

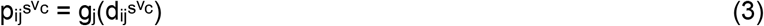

where g_j_(·) is the calibration function for feature j. The XGBoost branch produced a probability directly from the same input vector [27]:

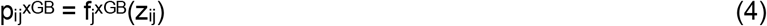

where f_j_^xGB^(·) denotes the boosted-tree ensemble for feature j. The three probabilities were then combined by equal-weight soft voting:

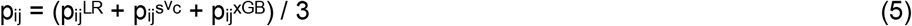

For patient k, visit-level probabilities were averaged across all visits V_k_ to obtain one patient-level probability per feature:

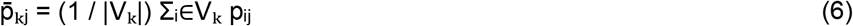

The resulting 44-dimensional patient vector 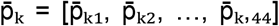 was used as input to a second-stage XGBoost stacker:

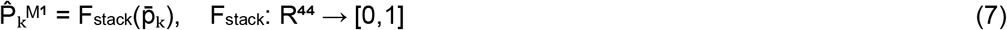

If feature j was completely absent for patient k, its value was replaced by the mean out-of-fold (OOF) training probability for that feature:

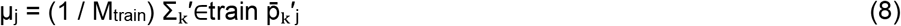

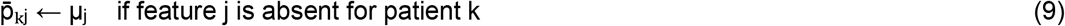

where M_train_ is the number of patients in training data. The final M1 class label was obtained using a decision threshold of 0.5:

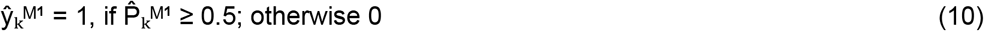

For the LR and calibrated SVC branches, inputs were standardised within each training fold and the same transformation was applied to the corresponding validation fold, whereas XGBoost used unscaled inputs. The final selected M1 configuration used the baseline trio at the feature level with an XGBoost stacker, while other M1 variants are summarised in Table 1.

**Table 1.** M1 mini-model variants. Overview of the 12 M1 feature-level ensemble variants evaluated in this study. Variants V0 to V3 compare alternative per-feature learners, including Logistic Regression (LR), Support Vector Machine (SVM) with calibration, and Extreme Gradient Boosting (XGB), used either individually or in combination. Variants V4 to V7 retain the baseline feature-level ensemble but modify the stacking model, including Logistic Regression, Extreme Gradient Boosting, Random Forest (RF), and Extra Trees (ET; Extremely Randomized Trees). Variants V8 to V11 assess alternative final-combination strategies, including area under the receiver operating characteristic curve (AUC)-weighted soft voting, Least Absolute Shrinkage and Selection Operator (LASSO) stacking, Elastic Net stacking, and Principal Component Analysis (PCA) followed by Logistic Regression. Predictions are generated at the visit level and then aggregated to the patient (PATNO) level before evaluation.

| Variant | Shorthand name | Algorithms used | Purpose |
| --- | --- | --- | --- |
| V0 | v0_baseline | LR + Calibrated SVM<br>+ XGB → LR stacker | Main baseline ensemble. Combines three per-feature learners, then uses logistic regression as the meta-model. |
| V1 | v1_lr_only | LR only → LR stacker | Tests whether simple logistic regression alone is enough at feature level. |
| V2 | v2_lr_xgb | LR + XGB → LR stacker | Removes SVM and keeps a lighter two-model per-feature ensemble. |
| V3 | v3_svm_only | Calibrated linear SVM → LR stacker | Isolates the SVM branch to measure its standalone contribution. |
| V4 | v4_ridge_stack | Baseline trio → Ridge-style LR stacker | Same feature ensemble as V0, but stacker uses a less strongly regularised L2 logistic regression. |
| V5 | v5_xgb_stack | Baseline trio → XGBoost stacker | Replaces linear stacking with a boosted-tree meta-learner. |
| V6 | v6_rf_stack | Baseline trio → Random Forest stacker | Uses a bagged tree ensemble as the meta-model to capture nonlinear interactions. |
| V7 | v7_et_stack | Baseline trio → Extra Trees stacker | Uses a more randomised tree ensemble as the meta-model. |
| V8 | v8_auc_weighted | Baseline trio → AUC-weighted soft vote | No learned stacker. Final prediction is a weighted average based on each feature's OOF AUC. |
| V9 | v9_lasso | Baseline trio → LASSO LR stacker | Uses L1-penalised logistic regression to shrink weak feature contributions to zero. |
| V10 | v10_elasticnet | Baseline trio → Elastic Net LR stacker | Uses a mixed L1 + L2 logistic stacker, useful when feature probabilities are correlated. |
| V11 | v11_pca_lr | Baseline trio → PCA + LR | Reduces the stacked feature space with PCA, then fits logistic regression on the components. |

#### 2.3.2. Development of M2: LightGBM-Only Global Model

M2 applied a single LightGBM classifier to the full multivariate feature space, thereby modelling all 44 clinical variables jointly together with age and sex. For each visit i, the input vector was defined as x_i_ = [x_i1_, x_i2_, …, x_i44_, a_i_, s_i_]^T^ ∈ ^46^, where x_ij_ denotes the value of feature j at visit i. Missing values were imputed using training-set column means:

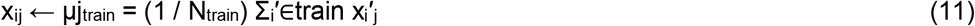

where μ j _train_is the mean of feature j in the training partition and N_train_ is the number of training visit rows. LightGBM then generated an additive score by summing the outputs of T boosted trees [28]:

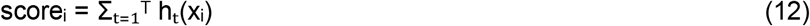

where h_t_(·) denotes the t-th tree. This score was converted to a visit-level PD probability using the sigmoid function:

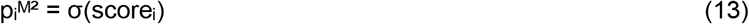

Because multiple visits could belong to the same patient, visit-level probabilities were averaged to obtain a patient-level M2 probability:

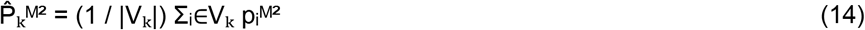

The final M2 class prediction was then defined as:

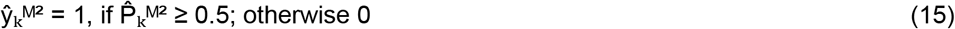

Unlike M1, which models each feature separately, M2 captures cross-feature interactions directly within a single global model. The final selected M2 configuration corresponded to the LightGBM-only model, while the remaining M2 variants are provided in Table 2.

**Table 2.** M2 mini-model variants. Overview of the 12 M2 global-ensemble variants evaluated in this study. Variants V0 to V3 compare the baseline ensemble with single-model baselines based on Extreme Gradient Boosting (XGB), Random Forest (RF), and Light Gradient Boosting Machine (LGB; LightGBM). Variants V4 to V7 assess reduced two-model combinations and a substitution of Extra Trees (ET; Extremely Randomized Trees) for one of the baseline learners. Variants V8 to V11 evaluate enhanced ensemble strategies, including replacement of RF with ET, use of a tuned XGB configuration, an out-of-fold (OOF) Logistic Regression (LR) stacker trained on base-model predictions, and an all-four-model soft-voting ensemble. Predictions are generated at the visit level and then aggregated to the patient (PATNO) level before evaluation.

| Variant | Shorthand name | Algorithms used | Purpose |
| --- | --- | --- | --- |
| V0 | v0_baseline | XGB + RF + LGB<br>equal soft vote | Main baseline global ensemble. Averages probabilities from Extreme Gradient Boosting (XGB), Random Forest (RF), and Light Gradient Boosting Machine (LGB / LightGBM). |
| V1 | v1_xgb_only | XGBoost only | Single-model baseline used to isolate the contribution of XGB alone. |
| V2 | v2_rf_only | Random Forest only | Single-model baseline used to test RF alone. |
| V3 | v3_lgb_only | LightGBM only | Single-model baseline used to test LGB / LightGBM alone. |
| V4 | v4_xgb_lgb | XGB + LGB soft vote | Two-model ensemble that removes RF to test whether the two boosting models work better together. |
| V5 | v5_xgb_rf | XGB + RF soft vote | Two-model ensemble that removes LGB, combining one boosting model with one bagging model. |
| V6 | v6_rf_lgb | RF + LGB soft vote | Two-model ensemble that removes XGB to test RF with LightGBM only. |
| V7 | v7_et_xgb | ET + XGB soft vote | Replaces RF and LGB with Extra Trees (ET; Extremely Randomized Trees) plus XGB, giving a more randomised tree-based pair. |
| V8 | v8_et_xgb_lgb | ET + XGB + LGB soft vote | Replaces RF in the baseline with ET, testing whether ET is a better tree-ensemble partner than RF. |
| V9 | v9_deep_xgb | Tuned XGB + RF + LGB soft vote | Keeps the baseline trio but uses a deeper/tuned XGB model, changing max_depth from 4 to 6 and n_estimators from 400 to 600. |
| V10 | v10_oof_stack | OOF LR stacker on XGB + RF + LGB | Uses out-of-fold (OOF) Logistic Regression (LR) as a meta-learner. Base-model predictions from XGB, RF, and LGB are generated under PATNO-level GroupKFold, then combined by LR instead of simple averaging. |
| V11 | v11_all4_ensemble | ET + XGB + RF + LGB equal soft vote | All-four ensemble. Adds ET to the baseline trio to test whether a broader multi-model average improves performance. |

#### 2.3.3 Development of M3: collaborative-filtering model

M3 was designed as a patient-similarity model using a CF framework [29, 30]. It operated on the patient-level matrix derived from the 50%-sparse dataset. For each patient k, repeated visits were collapsed into a single profile by averaging feature values across all visits V_k_:

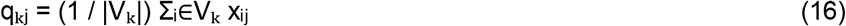

where q_kj_ denotes the patient-level value of feature _j_. The resulting patient vector was q_k_ = [q_k1_, q_k2_, …, q_k_,_44_, a_k_, s_k_]^T^, with missing entries imputed using training-set means. Cosine and Pearson similarities were computed on the imputed patient vectors, whereas the Euclidean branch used standardised vectors 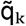. Cosine and Pearson similarities between patients k and l were defined as:

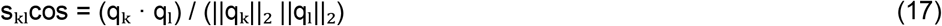

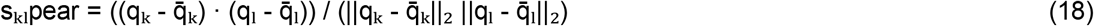

where 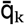denotes the mean-centred version of q_k_. For the Euclidean branch, pairwise distance and radial basis function (RBF) similarity were defined as:

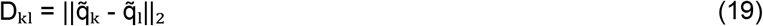

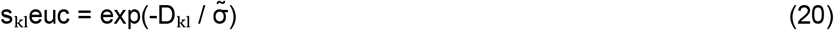

where 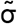 is the median pairwise distance among training patients. For each similarity metric m ∈{cos, pear, euc}, the K = 5 nearest neighbours of patient k were identified, and a similarity-weighted vote was computed as:

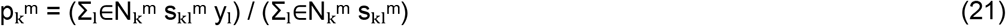

where N_k_^m^ denotes the neighbour set under metric m, and y_1_ is the known class label of neighbour l. The final M3 probability was obtained by averaging the three metric-specific probabilities:

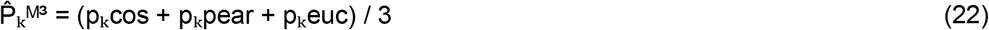

The final binary M3 prediction was defined as:

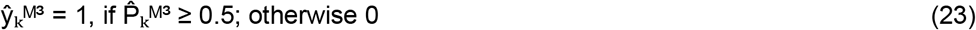

Thus, M3 complemented the supervised models by exploiting similarity structure among patient profiles rather than learning an explicit discriminative function. The final selected M3 configuration used the three-metric ensemble with K = 5, whereas other M3 variants are summarised in Table 3.

**Table 3.**
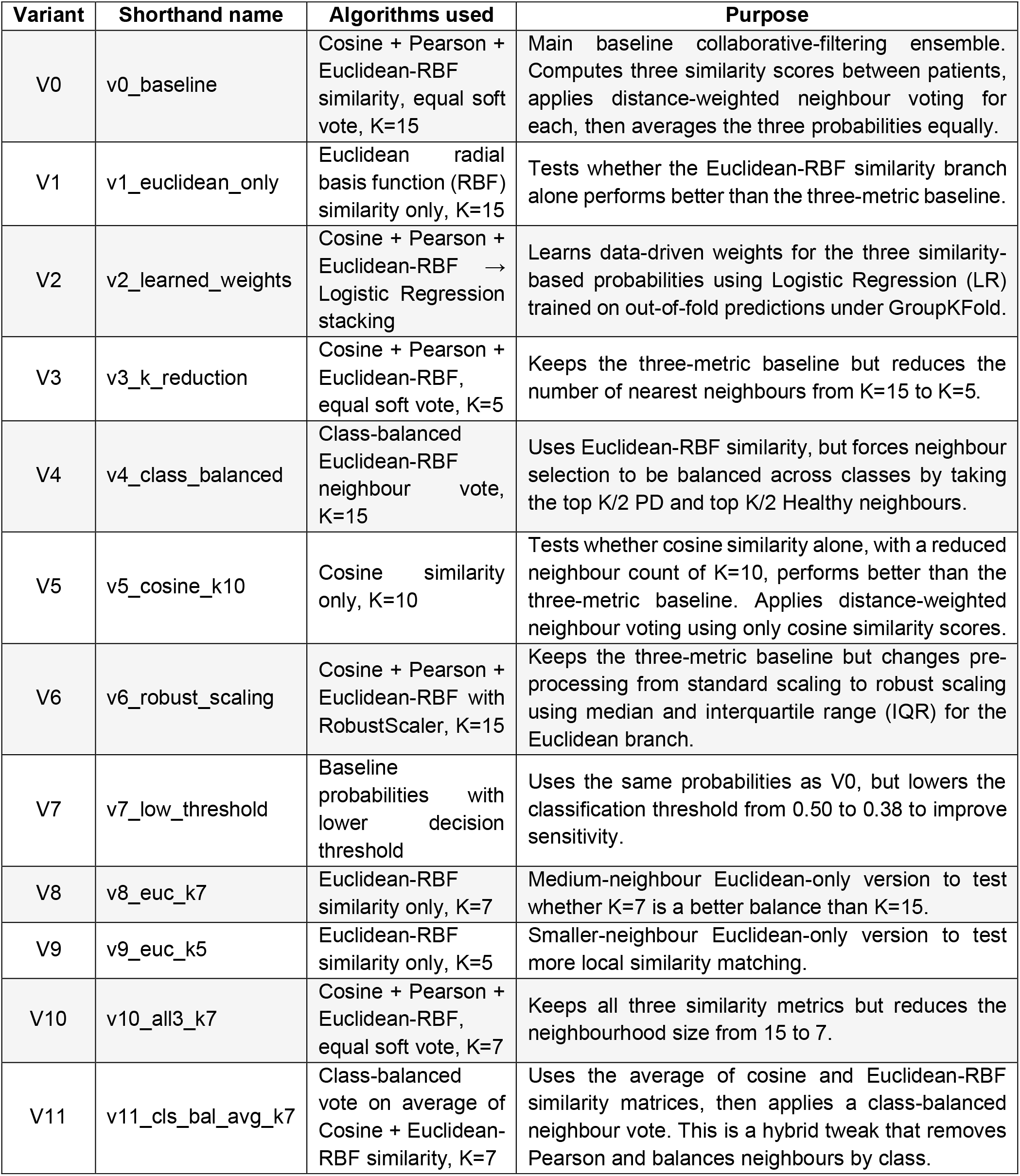
Overview of the 12 M3 collaborative-filtering variants evaluated in this study. Variants V0 to V2 compare the baseline ensemble of cosine similarity, Pearson correlation similarity, and Euclidean radial basis function (RBF) similarity with a Euclidean-only version and a Logistic Regression (LR)-stacked metric combiner trained on out-of-fold predictions. Variants V3 to V5 examine the effect of neighbourhood size reduction, class-balanced neighbour selection, and restricting similarity to cosine only at a reduced neighbourhood size (K=10).

Variants V6 to V11 assess additional refinements, including robust scaling using median and interquartile range (IQR), a reduced decision threshold, alternative Euclidean-only neighbourhood sizes, a three-metric ensemble with K = 7, and a hybrid class-balanced vote based on the average of cosine and Euclidean-RBF similarities. In all cases, the model operates at the patient (PATNO) level, using similarity-based neighbour voting to generate predicted probabilities.

### 2.4 Meta-Stacking and Final Ensemble

The final ensemble combined the patient-level outputs of M1, M2, and M3 through a Logistic Regression meta-learner. For each training patient k, grouped cross-validation generated three out-of-fold (OOF) probabilities, namely 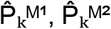and 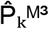, where each pro a ilit was o tained from a fold in which that patient was excluded from model training. These three values formed the meta-feature vector:

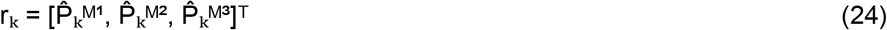

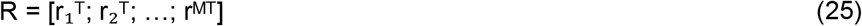

Stacking r_k_ across all M training patients produced the meta-feature matrix R of dimension M × 3, where M denotes the number of patients in the training set. A Logistic Regression meta-learner with balanced class weights was then fitted on R to estimate the final ensemble probability:

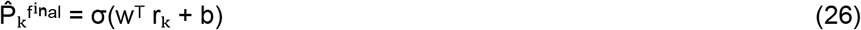

Here, w = [w_1_, w_2_, w_3_]^T^ contains the learned weights assigned to M, M, and M, is the intercept, and σ(·) is the sigmoid function. The final class label was obtained using a decision threshold of 0.5:

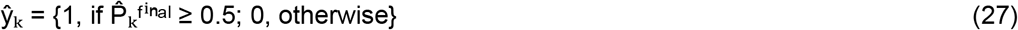

After training the meta-learner on OOF predictions, the three sub-models were retrained on the complete training set, and their patient-level probabilities were passed through the fixed meta-learner to generate final predictions for unseen patients. To provide an interpretable reliability measure, a confidence score was also computed from the final ensemble output. First, the probability margin from the decision boundary was defined as:

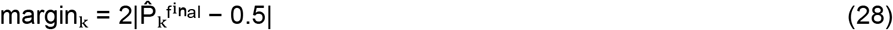

Here, margin_k_ ranges from 0 to 1 and increases as the predicted probability moves away from 0.5. Second, model consensus was defined as the proportion of sub-model predictions that agreed with the final class label:

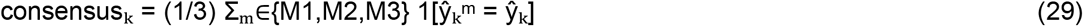

Here, ŷ_k_^m^ denotes the inar prediction of su model m for patient k, and [·] is the indicator function. The final confidence score combined both components as:

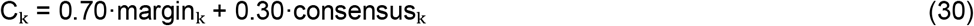

This confidence score was used only as an interpretive indicator, with higher values reflecting more decisive and more concordant predictions across the ensemble. Other than this auxiliary score, the final classification depended solely on the stacked probability defined in Eq. (26).

### 2.5 Interpretability Analysis

Model interpretability was assessed at two levels using SHapley Additive exPlanations (SHAP) [31], a game-theoretic framework that assigns each feature a contribution value reflecting its marginal impact on the model output. At the feature level, a TreeExplainer was applied to the M2 LightGBM model on patient-aggregated test data (visit rows averaged per PATNO) to quantify the mean absolute SHAP value of each of the 44 clinical features, thereby identifying which raw clinical domains most strongly influenced M2’s predictions. At the sub-model level, a LinearExplainer was applied to the Logistic Regression meta model, using the three-column OOF probability matrix as input, to quantify how much each sub-model (M1, M2, M3) contributed to the final ensemble output on average. These two analytical levels together provide interpretability both at the clinical feature granularity and at the ensemble architecture level, supporting transparency for clinical stakeholders.

### 2.6 Tool Development: PD-INSPECT

The trained ensemble was deployed as an interactive web application, designated PD-INSPECT, developed using the Flask microframework [32] and hosted on a server-constrained environment. The application accepts a minimum of 10 user-specified clinical feature scores alongside age and sex, and returns a risk classification, a final ensemble probability, and a structured confidence score. Thread allocation was restricted to a single thread per computational library (OpenMP, MKL, OpenBLAS, NumExpr) at the environment level to prevent out-of-memory errors on the deployment server.

The confidence score was computed as a weighted combination of two components: the probability margin, defined as twice the absolute deviation of the final probability from the decision boundary of 0.5 (i.e., 2 x |P_final -0.5|), capturing how decisively the ensemble classified the patient; and the model consensus, defined as the proportion of sub-models (M1, M2, M3) whose individual probabilities agreed with the final class prediction. The two components were combined as: Confidence = 0.70 x Probability Margin + 0.30 x Model Consensus. The resulting composite score was mapped to four interpretive bands: High Confidence (score >= 0.80), Moderate Confidence (score >= 0.55), Low Confidence (score >= 0.35), and Very Low Confidence (score < 0.35) [Table 4]. A downloadable PDF report, generated using the ReportLab library [33], provided a structured summary of the patient’s input, the risk classification, per-model probability breakdown, and the confidence assessment. The application retrieved feature names from the M3 CF bundle and applied neutral imputation for any features not supplied by the user, ensuring that partial clinical profiles could still yield a prediction.

**Table 4.** Interpretation of confidence levels based on the composite confidence score C_k_.

| Confidence Level | Condition on $C_k$ | Meaning |
| --- | --- | --- |
| High | $C_k \geq 0.80$ | Model is decisive and all/most sub-models agree. |
| Moderate | $0.55 \leq C_k < 0.80$ | Reasonable certainty; some uncertainty remains. |
| Low | $0.35 \leq C_k < 0.55$ | Prediction near the boundary or sub-models disagree. |
| Very Low | $C_k < 0.35$ | High uncertainty; interpret with caution. |

#### Confidence levels

### 2.8 Performance Metrics

Model performance was evaluated on a held-out test set at the patient level (PATNO), with visit-level probabilities averaged per patient before computing any metric. The primary metric was the Area Under the Receiver Operating Characteristic Curve (AUROC), with a 95% confidence interval estimated by 1,000-iteration stratified bootstrap resampling. Secondary metrics included sensitivity, specificity, F1 score, balanced accuracy, precision, and the Brier score, the latter measuring probability calibration as the mean squared difference between predicted probability and true binary label. All metrics were computed at a decision threshold of 0.5. The Brier score was selected as a calibration indicator because well-calibrated probabilities are critical for clinical confidence scoring; a lower Brier score indicates that the model’s predicted probabilities more accurately reflect true class frequencies [34]. All the important libraries used in analysis have been listed in Table S2.

## 3. Results

### 3.1 Variant Selection for Each Sub-Model

Each of the three sub-models was subjected to a structured evaluation of 12 architectural variants before a final configuration was selected. The rationale, variant descriptions, and supporting confusion matrices for all 36 variants are provided in Supplementary Figures S1-S3; key variant results are summarised in Tables 4-6 below.

#### 3.1.1 M1 Feature-Level Ensemble Variants

Across the 12 M1 variants, the choice of stacker proved more consequential than the base learner composition. Variants employing the Baseline Trio as base learners (V0, V4 through V7, V11) consistently outperformed single-algorithm alternatives such as LR-only (V1; AUC = 0.858) and SVM-only (V3; AUC = 0.916), confirming that ensemble diversity at the per-feature level contributes positively to discrimination. Among stacker types evaluated with the Baseline Trio as the shared base learner, replacing the regularised LR stacker (V0; AUC = 0.927, balanced accuracy = 0.877) with an XGBoost stacker yielded the strongest discrimination (V5; AUC = 0.983, sensitivity = 0.971, specificity = 0.925, balanced accuracy = 0.948). The Random Forest and ExtraTrees stackers (V6 and V7) achieved comparably high AUC values (0.988 and 0.988, respectively) but at lower specificity (0.875 and 0.850), indicating a sensitivity-specificity imbalance less desirable for clinical screening. The AUC-Weighted Soft Vote variant (V8) achieved unit sensitivity but zero specificity, rendering it unsuitable. V5 was therefore selected as the final M1 configuration on account of its superior balanced accuracy and AUC. Performance metrics of the M1 mini-model variants are summarised in Table 5, while the corresponding confusion matrices are provided in Figure S1.

**Table 5.** M1 feature-ensemble variant comparison. All 12 variants evaluated on the held-out test set at the PATNO level. Reported metrics include area under the receiver operating characteristic curve (AUC), sensitivity (Sens.; true positive rate), specificity (Spec.; true negative rate), F1 score (harmonic mean of precision and recall), and balanced accuracy (Bal. Acc.; mean of sensitivity and specificity). The selected configuration (V5) was chosen based on the highest AUC and balanced accuracy. Full confusion matrices are provided in Supplementary Figure S1.

| Variant | AUC | Sens. | Spec. | F1 | Bal. Acc. | Selection |
| --- | --- | --- | --- | --- | --- | --- |
| V0 | 0.9273 | 0.9296 | 0.825 | 0.9395 | 0.8773 |  |
| V1 | 0.8579 | 0.8239 | 0.725 | 0.8667 | 0.7745 |  |
| V2 | 0.9407 | 0.9296 | 0.825 | 0.9395 | 0.8773 |  |
| V3 | 0.9164 | 0.9155 | 0.775 | 0.9253 | 0.8452 |  |
| V4 | 0.9694 | 0.9648 | 0.825 | 0.958 | 0.8949 |  |
| **V5** | **0.9838** | **0.9718** | **0.925** | **0.9753** | **0.9484** | ✓ |
| V6 | 0.9887 | 0.9718 | 0.875 | 0.9684 | 0.9234 |  |
| V7 | 0.9883 | 0.9718 | 0.85 | 0.965 | 0.9109 |  |
| V8 | 0.9231 | 1 | 0 | 0.8765 | 0.5 |  |
| V9 | 0.8269 | 0.8662 | 0.65 | 0.8817 | 0.7581 |  |
| V10 | 0.9232 | 0.9225 | 0.825 | 0.9357 | 0.8738 |  |
| V11 | 0.9695 | 0.9648 | 0.825 | 0.958 | 0.8949 |  |

#### 3.1.2 M2 LightGBM-Only Model Variants

The 12 M2 variants demonstrated uniformly high sensitivity across all configurations (1.000 in all 12 variants), as a consequence of scale_pos_weight-based class balancing or equivalent weighting strategies, which ensured near-complete capture of PD cases. Accordingly, specificity served as the primary discriminator among variants. The LightGBM-only configuration (V3) achieved a specificity of 0.800 and balanced accuracy of 0.900 among the single-model variants, outperforming random forest-inclusive combinations: V2 (RandomForest Only) dropped to specificity 0.400, while mixed ensembles involving random forest (V5, V6) showed intermediate specificity (0.725 and 0.675, respectively). The XGBoost-only variant (V1) achieved the highest raw specificity of 0.825, but at the cost of modelling simplicity and compatibility with the inference pipeline, which requires explicit missing value imputation. The OOF LR stacking variant (V10) recovered a specificity of 0.800 comparable to V3, but introduced additional complexity through the stacking layer. V3 was therefore designated the final M2 configuration, given its strong balance of sensitivity and specificity, its compatibility with the production imputation pipeline, and its computationally efficient gradient boosting framework [28]. Performance metrics of the M2 mini-model variants are summarised in Table 6, while the corresponding confusion matrices are provided in Figure S2.

**Table 6.** M2 global-ensemble variant comparison. All 12 variants evaluated on the held-out test set at the PATNO level. Given uniformly perfect sensitivity, specificity and balanced accuracy serve as primary discriminators. The selected configuration (V3) achieved a strong balance of sensitivity and specificity, with LightGBM chosen as the final M2 configuration on account of its computational efficiency, compatibility with the inference pipeline, and competitive balanced accuracy. Full confusion matrices are provided in Supplementary Figure S2.

| Variant | AUC | Sens. | Spec. | F1 | Bal. Acc. | Selection |
| --- | --- | --- | --- | --- | --- | --- |
| V0 | 0.9755 | 1 | 0.75 | 0.966 | 0.875 |  |
| V1 | 0.9776 | 1 | 0.775 | 0.9693 | 0.8875 |  |
| V2 | 0.9723 | 1 | 0.4 | 0.9221 | 0.7 |  |
| **V3** | **0.9706** | **1** | **0.8** | **0.9726** | **0.9** | ✓ |
| V4 | 0.9755 | 1 | 0.775 | 0.9693 | 0.8875 |  |
| V5 | 0.9778 | 1 | 0.725 | 0.9627 | 0.8625 |  |
| V6 | 0.9734 | 1 | 0.675 | 0.9562 | 0.8375 |  |
| V7 | 0.9783 | 1 | 0.7 | 0.9595 | 0.85 |  |
| V8 | 0.9757 | 1 | 0.75 | 0.966 | 0.875 |  |
| V9 | 0.9761 | 1 | 0.7 | 0.9595 | 0.85 |  |
| V10 | 0.9759 | 1 | 0.775 | 0.9693 | 0.8875 |  |
| V11 | 0.9761 | 1 | 0.675 | 0.9562 | 0.8375 |  |

#### 3.1.3 M3 Collaborative Filtering Variants

Among the 12 M3 variants, sensitivity remained consistently high across configurations, while specificity varied substantially, constituting the primary basis for selection. The baseline variant (V0; K = 15, combined metrics) achieved high sensitivity (0.979) but limited specificity (0.500). Reducing the neighbourhood to K = 5 while retaining the three-metric combination (V3; cosine + Pearson + Euclidean RBF) improved specificity to 0.625 while maintaining sensitivity at 0.965, yielding the highest balanced accuracy (0.795) of any M3 variant. Learned weighting via LR stacking (V2) maximised sensitivity (0.993) but further deteriorated specificity (0.375), rendering it clinically unsuitable. Euclidean-only variants (V1, V8, V9) showed lower AUC values (0.854 to 0.902) compared to the combined-metric variants, supporting the complementarity of multiple similarity measures. V3 was selected as the final M3 configuration on account of its superior specificity-sensitivity balance. Performance metrics of the M3 mini-model variants are summarised in Table 7, while the corresponding confusion matrices are provided in Figure S3. Confusion matrices comparison of selected M1, M2, and M3 Models is shown in Figure 2.

**Table 7.** M3 collaborative filtering variant comparison. All 12 variants evaluated on the held-out test set at the PATNO level. Specificity served as the primary discriminator given consistently high sensitivity. The selected configuration (V3) achieved the best-balanced accuracy among all variants. Full confusion matrices are provided in Supplementary Figure S3.

| Variant | AUC | Sens. | Spec. | F1 | Bal. Acc. | Selection |
| --- | --- | --- | --- | --- | --- | --- |
| V0 | 0.940 | 0.979 | 0.500 | 0.924 | 0.739 |  |
| V1 | 0.902 | 0.944 | 0.600 | 0.918 | 0.772 |  |
| V2 | 0.939 | 0.993 | 0.375 | 0.916 | 0.684 |  |
| **V3** | **0.921** | **0.965** | **0.625** | **0.932** | **0.795** | ✓ |
| V4 | 0.888 | 0.916 | 0.625 | 0.906 | 0.770 |  |
| V5 | 0.894 | 0.965 | 0.5 | 0.916 | 0.732 |  |
| V6 | 0.892 | 0.972 | 0.325 | 0.899 | 0.648 |  |
| V7 | 0.940 | 1.000 | 0.275 | 0.907 | 0.638 |  |
| V8 | 0.869 | 0.916 | 0.650 | 0.909 | 0.783 |  |
| V9 | 0.854 | 0.916 | 0.600 | 0.903 | 0.758 |  |
| V10 | 0.926 | 0.965 | 0.575 | 0.926 | 0.770 |  |
| V11 | 0.863 | 0.887 | 0.625 | 0.891 | 0.756 |  |

**Fig. 2.**
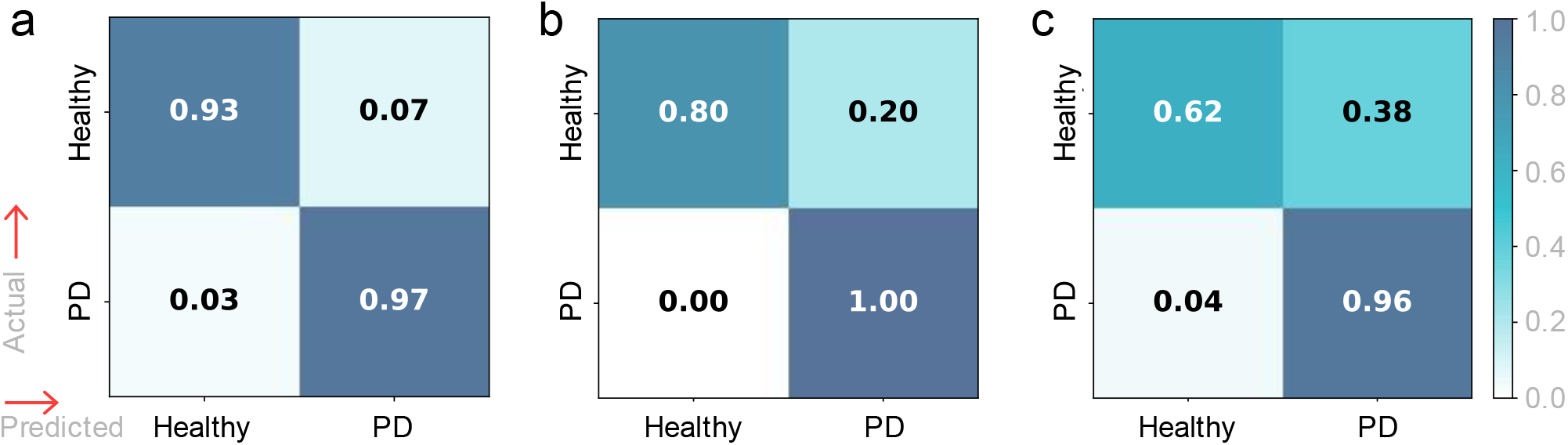
(a) Final M1 ensemble (V5; XGBoost stacker) demonstrating balanced performance with sensitivity 0.972 and specificity 0.925. (b) Final M2 model (LightGBM-only, V3) achieving perfect sensitivity (1.000) and the highest specificity (0.800) among M2 variants. (c) Final M3 configuration (K=5 collaborative filtering, V3) maintaining high sensitivity (0.965) but comparatively lower specificity (0.625). Cell values represent counts of Healthy and PD classifications at the PATNO level. Differences across models are primarily driven by specificity, with M2 providing the strongest overall class separation.

### 3.2 Final Ensemble Performance

Meta-stacking of the three sub-models through a Logistic Regression classifier demonstrated strong and balanced performance across all evaluation metrics. On the held-out test set of 182 patients (142 PD, 40 healthy), the final ensemble achieved an AUC of 0.9937 (95% CI [0.984, 0.999]), correctly classifying 138 of 142 PD patients (sensitivity = 0.9718) and 37 of 40 healthy controls (specificity = 0.925). The balanced accuracy of 0.9484 exceeded that of every individual sub-model, confirming that meta-stacking improved overall class balance beyond what any constituent model achieved alone. The F1 score of 0.9753 and precision of 0.9787 indicate high positive predictive value, implying that a PD classification from the ensemble carries substantial clinical reliability. The Brier score of 0.0314 was the lowest among all models evaluated, reflecting superior probability calibration in the final ensemble relative to the individual sub-models.

The observed improvement in specificity from individual sub-models to the final ensemble warrants particular attention. While M2 and M3 individually showed relatively low specificity (0.8 and 0.625, respectively), the meta learner effectively learned to down-weight their high-sensitivity signals where they diverged from M1’s more conservative per-feature evidence. The Logistic Regression meta model explicitly learned a weighting of the three sub-model probabilities; the dominance of M1’s contribution in the final prediction, as confirmed by subsequent SHAP analysis (Section 3.4), suggests that the feature-level ensemble provided the most reliable discrimination signal for the meta learner to anchor upon, while M2 and M3 contributed complementary information that reduced overall misclassification. Performance metrics of the selected configurations for the three sub-models (M1, M2, and M3) and the final meta-stacked ensemble are summarised in Table 8, while the corresponding AUROC plot is provided in Figure S4. Performance of the final meta-stacked ensemble model shown in figure 3.

**Table 8.** Comparative performance of the selected sub-models and the final ensemble. Performance metrics for the chosen configurations of the three sub-models (M1 feature-level ensemble, M2 LightGBM model, and M3 collaborative filtering model) and the final meta-stacked ensemble evaluated on the held-out PATNO-level test set (n = 182; 142 PD, 40 healthy controls). Metrics reported include area under the ROC curve (AUC), sensitivity, specificity, precision, F1 score, balanced accuracy, and Brier score. The ensemble model demonstrates improved class balance and calibration relative to individual sub-models, achieving the highest balanced accuracy and the lowest Brier scores among all evaluated models.

| Model | AUC | Sens. | Spec. | Precision | F1 | Bal. Acc. | Brier Score |
| --- | --- | --- | --- | --- | --- | --- | --- |
| M1 | 0.9838 | 0.9718 | **0.925** | **0.9787** | **0.9753** | **0.9484** | 0.034 |
| M2 | 0.9706 | **1** | 0.8 | 0.9467 | 0.9726 | 0.9 | 0.0506 |
| M3 | 0.9205 | 0.9648 | 0.625 | 0.9013 | 0.932 | 0.7949 | 0.0902 |
| *Final Ensemble* | **0.9937** | 0.9718 | **0.925** | **0.9787** | **0.9753** | **0.9484** | **0.0314** |

**Fig. 3.**
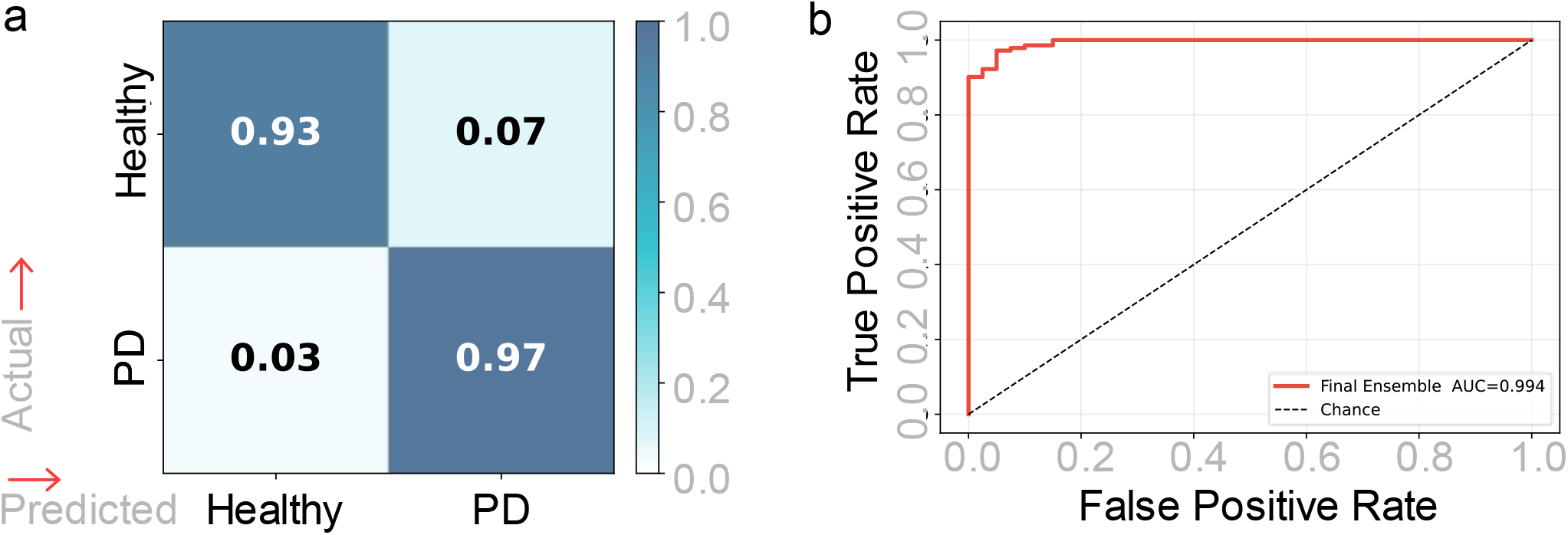
Performance of the final meta-stacked ensemble model. (a) Confusion matrix of the final ensemble evaluated on the held-out PATNO-level test set (n = 182; 142 PD, 40 healthy controls). The model correctly classified 138 of 142 PD patients and 37 of 40 healthy controls, corresponding to a sensitivity of 0.9718 and specificity of 0.925. (b) Receiver operating characteristic (ROC) curve of the final ensemble, showing strong discriminative ability with an AUC of 0.994. The dashed diagonal line indicates chance-level performance. Together, these results demonstrate the ensem le’s strong diagnostic discrimination and alanced classification performance.

### 3.3 Feature Interpretability via SHAP Analysis

At the raw clinical feature level, SHAP analysis of the M2 LightGBM global model (TreeExplainer applied to patient-aggregated test data) identified olfactory function (Smell) as the single most influential feature, with a mean absolute SHAP value of 1.14 followed by gastrointestinal symptoms (1.05) and cardiovascular autonomic features (0.54). Top ten of them are shown in Figure 4.a. The prominence of these non-motor and autonomic features is consistent with the established prodromal and early-stage profile of PD, in which olfactory loss and autonomic dysfunction frequently precede or accompany motor symptom onset [14, 35]. All SHAP values are reported as mean absolute values rounded to two decimal places for clarity.

**Fig. 4.**
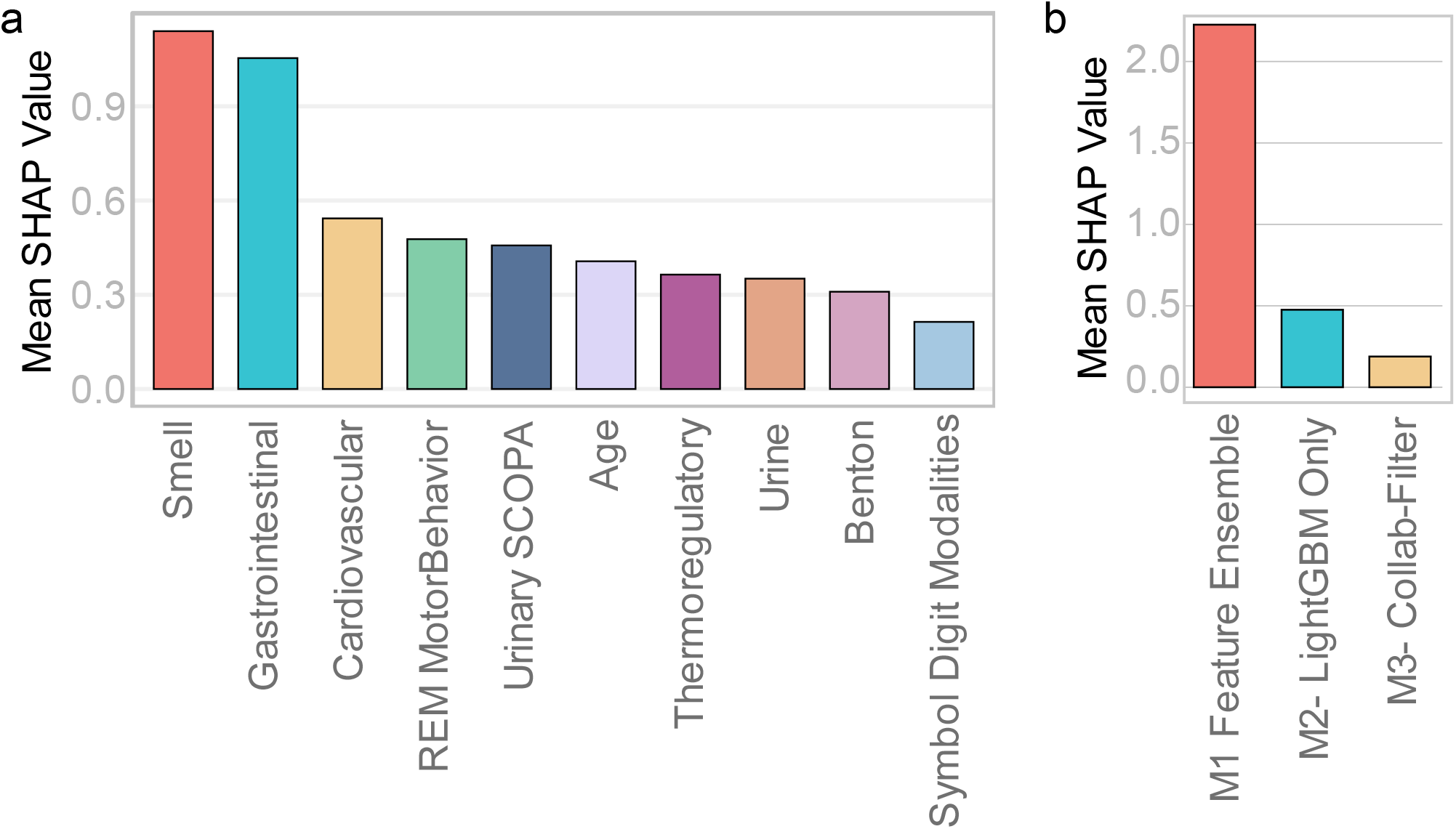
SHAP-based interpretability of the stacked ensemble framework. (a) Mean absolute SHAP values of the top ten clinical features derived from the M2 LightGBM global model, illustrating their relative contribution to PD prediction. Olfactory dysfunction (Smell) shows the strongest influence, followed by gastrointestinal and cardiovascular autonomic features, with additional contributions from REM sleep behaviour, urinary symptoms, thermoregulatory measures, and cognitive tests (Benton and Symbol Digit Modalities). (b) Mean absolute SHAP contribution of each sub-model to the final meta-stacked ensemble prediction, computed using a LinearExplainer on the LR meta-learner. The feature-level ensemble (M1) exerts the dominant influence on the final prediction, while the LightGBM global model (M2) and the collaborative filtering module (M3) provide complementary but comparatively smaller contributions. These results highlight the central role of feature-level modelling in the ensemble while demonstrating the complementary signals captured by the other sub-models. Each SHAP value is computed at the individual-sample level; for global feature importance, the bar plots summarise these local contributions by reporting the mean absolute SHAP value across all samples for each feature.

SHAP analysis at the sub-model level (LinearExplainer applied to the meta logistic regression) confirmed that M1 Feature-Ensemble exerted the strongest influence on the final ensemble prediction, with a mean absolute SHAP value of 2.226, substantially exceeding M2 LightGBM-Only (0.476) and M3 Collaborative Filtering (0.188; Figure 4b). This ordering aligns with the individual AUC-specificity profile of each sub-model: M1’s high specificity made its probability estimates more discriminative at the meta level, and the meta learner assigned it commensurately greater weight.

### 3.4 Web Application for Clinical feasibility

The PD-INSPECT system was deployed as a web-based application allowing input of age, sex, and NMS. A minimum of 10 features is recommended for reliable prediction. The system provides probability outputs, confidence levels, and model-wise contributions. Outputs include downloadable PDF reports and a debug interface for validation of model integrity and reproducibility (Figure S5). The application is accessible online and does not require local installation. The tool can be accessed https://pdinspect.pythonanywhere.com. For transparency, the interface also provides model-wise probability outputs (M1, M2, and M3) and their contribution toward the final ensemble probability. The report includes a confidence band derived from probability margin and sub-model agreement, and a summary of which features were provided by the user. Representative examples of downloadable reports generated by the application for a healthy/low-risk prediction is provided in Supplementary Figure S5; all displayed demographic details (name, age, and sex) are fictional placeholder values included solely for interface demonstration purposes and do not correspond to real individuals.

## 4. Discussion

This study demonstrates the feasibility of using NMS as predictors of PD risk. SHAP analysis showed that olfactory, gastrointestinal, and urinary NMS were the strongest predictors, which aligns with known prodromal features of PD. Olfactory dysfunction was the strongest feature, consistent with early PD pathology [36]. Gastrointestinal NMS also showed high importance, supporting the gut-brain hypothesis [37]. Urinary and other autonomic features further indicate early autonomic involvement [10]. Cognitive measures such as Symbol Digit and Benton tests were also important, reflecting early cognitive decline in PD [38]. Agreement between SHAP and M1 feature ranking suggests robustness of these findings.

From a clinical perspective, the study shows that PD risk assessment with acceptable accuracy can be done using only NMS without imaging or molecular biomarkers. This makes the approach practical for screening. The achieved sensitivity and specificity indicate usefulness as a first-level screening tool. If trained on more data, in low-income countries, where specialist access is limited, such a tool can help in early screening [3]. The observed improvement in specificity is important for screening-oriented use cases, because it reduces false-positive risk flags in healthy controls while maintaining high sensitivity.

In this study, we developed and validated PD-INSPECT, a stacked ensemble framework for PD risk prediction using NMS data from the PPMI cohort. The final model achieved AUC of 0.994, sensitivity of 0.972, and specificity of 0.925 on the test set. This reflects a clear improvement in specificity compared to individual sub-models. In practical terms, the final ensemble reduces overconfident single-model behaviour by integrating calibrated sub-model probabilities through stacking [22, 34]. The main hypothesis was that combining different modelling strategies would improve performance over single models. The results support this. M1 provided strong overall balance, but its feature-wise modelling may underutilize cross-feature interactions captured by global models. M2 achieved the highest sensitivity but had lower specificity. The inclusion of CF is relatively novel in PD prediction. Although M3 had lower specificity, it captured patient similarity patterns that complemented supervised models [23]. The meta-learner corrected this imbalance by assigning higher weight to M1, which provided better-calibrated probabilities. This led to improved overall balance between sensitivity and specificity. The final model also showed highest AUC, balanced accuracy, and lowest Brier score, indicating good probability calibration, which is important for clinical decision support [22, 34].

Compared to previous studies, the performance is favourable. Earlier work using clinical data reported accuracy in the range of 70-80% [18]. The improvement in our model is likely due to stacked modelling and strict patient-level validation. Studies using speech or gait features have reported higher AUC, but they depend on specialized data collection [17]. In contrast, this study uses only NMS from routine clinical assessments.

The study has several strengths. It uses a heterogeneous ensemble combining feature-level, global, and similarity-based models. Patient-level cross-validation avoids data leakage and improves reliability [25]. Systematic evaluation of multiple variants adds transparency. SHAP-based explainability improves interpretability and clinical trust [31]. The web-based implementation primarily serves as a proof of feasibility and can be expanded in future as additional data become available.

Future work should focus on external validation on diverse datasets. Incorporating longitudinal NMS progression can improve early detection [39]. Integration with imaging or biomarker data may further enhance performance. Prospective clinical studies are needed to establish real-world utility. Federated learning can support multi-centre deployment while preserving data privacy [40].

## 5. Limitations

Although this study achieved significantly higher accuracy, it has several limitations. First, all data are from a single cohort (PPMI), which mainly includes North American and European participants, so external validation on independent cohorts from other population with different PD characteristics, is required before any clinical claims. Second, the class imbalance (142 PD vs. 40 healthy, about 3.5:1) means the reported metrics should be interpreted carefully; weighting methods in M1 and M2 help but still introduce some bias toward PD prediction, which is only partly corrected by the meta-stacker. Third, M3 shows lower specificity due to limitations of memory-based CF on sparse clinical profiles, where fewer input features reduce the reliability of similarity estimates despite a minimum feature requirement. Fourth, the model treats all assessment timepoints equally and does not capture disease progression; adding temporal features like rate of change or visit order could improve early detection, as patients are currently treated as static profiles. Fifth, the confidence scoring formula (0.70 × margin + 0.30 × consensus) is empirically defined and not clinically validated, and the weighting is assumed rather than data-driven.

## 6. Conclusion

This study shows that NMS can serve as effective predictors of PD risk. Using a stacked ensemble framework, PD-INSPECT achieved strong performance on an independent test cohort. These findings indicate that NMS-based clinical and cognitive features carry substantial discriminative value for PD risk stratification. The three sub-models captured complementary aspects of the NMS profile, and their integration improved overall class balance and probability reliability compared with individual models. SHAP analysis further showed that olfactory, gastrointestinal, and autonomic symptoms were among the main drivers of prediction, supporting consistency with known PD-related non-motor patterns. The study also presents a web-based implementation of the model to support accessible and transparent output generation. However, the main contribution of this work lies in demonstrating the predictive value of NMS for PD risk assessment.

## Data Availability

All data produced are available online at GitHub

https://github.com/iam-zain/pd_inspect

https://pdinspect.pythonanywhere.com

## Statements and Declarations

## Acknowledgments

We wish to express our gratitude to UGC-SAP for their support to the Department of Biotechnology & Bioinformatics and the Bioinformatics Infrastructure Facility (BIF) at the School of Life Sciences, University of Hyderabad, India, for providing essential support and infrastructure. Additionally, we also acknowledge CSIR – HRDG, India, for granting a fellowship to Md Zainul Ali during his studies. Data used in the preparation of this article were o tained [on nd e ruar 0] from the Parkinson’s Progression Markers Initiative (PPMI) database (www.ppmi-info.org/access-dataspecimens/download-data), RRID:SCR 006431. This analysis used Tier 1 (open access) PPMI data ppmi-publication policy. For up-to-date information on the study, visit www.ppmi-info.org “PPMI – a public-private partnership – is funded by the Michael J. Fox Foundation for Parkinson’s esearch and funding partners, including [A complete list of PPMI funding partners is available on the PPMI we site].”

## AI-assisted editing declaration

All substantive intellectual content, analysis, and interpretations remain the original work of the authors. Portions of the manuscript text were revised and refined with the assistance of AI-based language tools, and any AI-generated suggestions were critically reviewed and approved by the author team prior to submission.

## Author contributions

M.Z.A. and P.S.D. conceptualized the research aims. M.Z.A. performed data curation, formal analysis, investigation, methodology development, software implementation, and visualization. P.S.D. contributed to funding acquisition, project administration, resources, and supervision. Both authors contributed to interpretation of the results. M.Z.A. wrote the initial drafts of the manuscript, and P.S.D. reviewed and edited the manuscript. All authors read and approved the final manuscript.

## Code availability

Scripts for the data analysis https://github.com/iam-zain/pd_inspect

Tool Weblink: https://pdinspect.pythonanywhere.com/

## Ethical considerations

Data used in the preparation of this article were obtained from the PPMI database, which is publicly available and deidentified. The PPMI study has obtained the necessary ethical approvals for the collection and use of the data, in accordance with institutional and regulatory guidelines. No additional ethical approval was required for this secondary data analysis.

