## Supplementary Data for "Interpretable Symptom-Based Machine Learning for Parkinson’s Disease Prediction: A Feasibility Study"

**Supplementary Tables**

**Table S1:** Clinical variables and assessment instruments used in the predictive modelling framework

| Sl. | Category | Clinical Domain | Feature / Test Name |
|---|---|---|---|
| 1 | Anxiety & Mood | Anxiety | MDS Anxiety |
| 2 | | Anxiety (State) | State-Trait Anxiety Inventory – State (STAIS) |
| 3 | | Anxiety (Trait) | State-Trait Anxiety Inventory – Trait (STAIA) |
| 4 | | Apathy | MDS Apathy |
| 5 | | Depression | Geriatric Depression Scale (Short Version) |
| 6 | | Depression | MDS Depression |
| 7 | Autonomic Dysfunction | Autonomic – Cardiovascular | SCOPA-AUT Cardiovascular |
| 8 | | Autonomic – Cardiovascular (Orthostatic) | MDS-UPDRS Light-headedness |
| 9 | | Autonomic – Gastrointestinal | SCOPA-AUT Gastrointestinal |
| 10 | | Autonomic – Gastrointestinal (Constipation) | MDS-UPDRS Constipation |
| 11 | | Autonomic – Pupillomotor | SCOPA-AUT Pupillomotor |
| 12 | | Autonomic – Thermoregulatory | SCOPA-AUT Thermoregulation |
| 13 | | Autonomic – Urinary | SCOPA-AUT Urinary |
| 14 | | Autonomic – Urinary (Symptom) | MDS-UPDRS Urinary Problems |
| 15 | Cognition | Cognition – Executive Function | Trail Making Test Part B |
| 16 | | Cognition – Global | Cognitive Categorization Assessment |
| 17 | | Cognition – Global | MDS Cognition |
| 18 | | Cognition – Global | Montreal Cognitive Assessment (MoCA) |
| 19 | | Cognition – Language | Modified Boston Naming Test |
| 20 | | Cognition – Language / Executive | Lexical Fluency |
| 21 | | Cognition – Memory (Learning) | Hopkins Verbal Learning Test |
| 22 | | Cognition – Memory (Recognition) | Hopkins Verbal Learning Test Recognition |
| 23 | | Cognition – Processing Speed | Symbol Digit Modalities Test |
| 24 | | Cognition – Processing Speed / Attention | Trail Making Test Part A |
| 25 | | Cognition – Semantic Memory | Semantic Fluency |
| 26 | | Cognition – Visuospatial / Executive | Clock Drawing Test |
| 27 | | Cognition – Visuospatial Perception | Benton Judgment of Line Orientation |
| 28 | | Cognition – Working Memory | Letter–Number Sequencing |
| 29 | Impulse Control | Impulse Control Behavior | Impulsive-Compulsive Behavior (QUIP) |
| 30 | | Impulse Control Disorder (Drug-related) | Impulsive ICD |
| 31 | | Impulse Control / Dopamine Dysregulation | MDS Dopamine Dysregulation Syndrome |
| 32 | Motor-associated NMS | Fatigue | MDS-UPDRS Fatigue |
| 33 | | Pain | MDS-UPDRS Pain |
| 34 | Psychosis | Psychosis (Hallucinations) | MDS Hallucination |
| 35 | Sleep & REM | Sleep – Daytime Dysfunction | MDS-UPDRS Daytime Sleepiness |
| 36 | | Sleep – Daytime Sleepiness | Epworth Sleepiness Scale |
| 37 | | Sleep – Night Disturbance | MDS-UPDRS Night Sleep Problems |
| 38 | | Sleep – REM (Awakening Disturbance) | REM Awake Problem |
| 39 | | Sleep – REM (Dream Disturbance) | REM Dream |
| 40 | | Sleep – REM (Dream Recall) | REM Awake Dream |
| 41 | | Sleep – REM (Movement Disorder) | REM Movement |
| 42 | Olfaction | Olfaction | University of Pennsylvania Smell Identification Test (UPSIT) |
| 43 | Sociodemographic | Sociodemographic | Handedness |
| 44 | | Sociodemographic | Years of Education |

**Table S1-** The table lists the 44 clinical features used as input variables for model development, together with the associated assessment tests or instruments from which they were derived. The variables span multiple clinical domains relevant to Parkinson’s disease, including neuropsychiatric symptoms (e.g., anxiety, depression, apathy), cognitive and neuropsychological assessments, sleep disturbances, autonomic dysfunction (gastrointestinal, urinary, cardiovascular, and thermoregulatory functions), olfactory assessment, and behavioral features. All measures were obtained from the Parkinson’s Progression Markers Initiative (PPMI) dataset and used as numerical inputs in the machine-learning models after preprocessing and sparsity filtering.

**Table S2:** Software libraries and package versions used in the PD-INSPECT framework.

| Name | Version |
|---|---|
| blinker | 1.9.0 |
| charset-normalizer | 3.4.4 |
| click | 8.3.1 |
| cloudpickle | 3.1.2 |
| colorama | 0.4.6 |
| Flask | 3.1.3 |
| itsdangerous | 2.2.0 |
| Jinja2 | 3.1.6 |
| joblib | 1.5.3 |
| MarkupSafe | 3.0.3 |
| numpy | 2.2.6 |
| packaging | 26.0 |
| pandas | 2.2.3 |
| pillow | 12.1.1 |
| python-dateutil | 2.9.0.post0 |
| pytz | 2025.2 |
| reportlab | 4.4.10 |
| scikit-learn | 1.7.2 |
| scipy | 1.15.3 |
| six | 1.17.0 |
| slicer | 0.0.8 |
| threadpoolctl | 3.6.0 |
| tqdm | 4.67.3 |
| typing_extensions | 4.15.0 |
| tzdata | 2025.3 |
| Werkzeug | 3.1.6 |
| xgboost | 2.1.4 |

**Table S2-** List of Python libraries and corresponding version numbers used for model development, evaluation, explainability analysis, web deployment, and report generation. The environment includes machine learning libraries (scikit-learn, xgboost, scipy), data processing tools (numpy, pandas), visualization and utility packages, and web framework components (Flask, Jinja2, Werkzeug). Reporting functionality was implemented using reportlab. All versions are specified to ensure computational reproducibility and facilitate environment replication.

#### Supplementary Figure

**Figure S1:** Systematic comparison of 12 M1 feature-ensemble variants on the held-out test set.

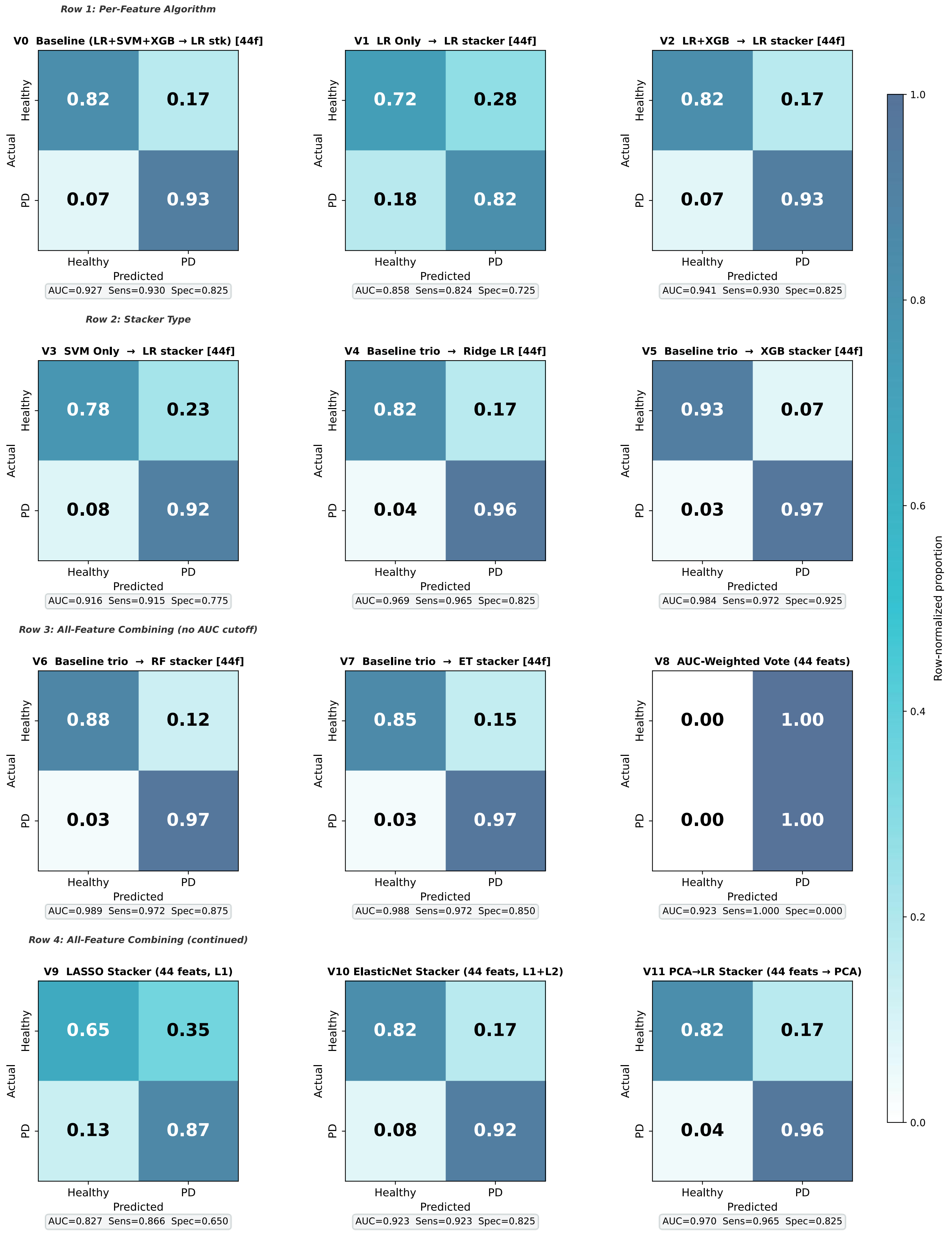

**Figure S1-** Confusion matrices for all predefined M1 configurations evaluated at the PATNO level (4×3 grid). Variants differ by base learner composition (LR, SVM, XGBoost), stacker type (Logistic Regression, Ridge, XGBoost, Random Forest, Extra Trees), and feature-selection threshold. The baseline trio with Logistic Regression stacker (V0) demonstrated balanced performance (AUC 0.899; sensitivity 0.908; specificity 0.800; balanced accuracy 0.854) and superior probability calibration, leading to its selection for meta-stacking. LR-only (V1) showed the weakest discrimination, highlighting the importance of ensemble diversity, while SVM-only (V3) achieved higher AUC (0.918), suggesting strong linear separability at the feature level. Replacing the LR stacker with XGBoost (V5) increased AUC (0.973) and sensitivity (0.972). Variations in feature-selection threshold (AUC > 0.60 and > 0.70) had minimal impact on overall performance, indicating limited contribution from lower-ranked features. Cell values represent counts of Healthy and PD classifications at a 0.5 decision threshold.

**Figure S2:** Systematic comparison of 12 M2 global-ensemble variants on the held-out test set.

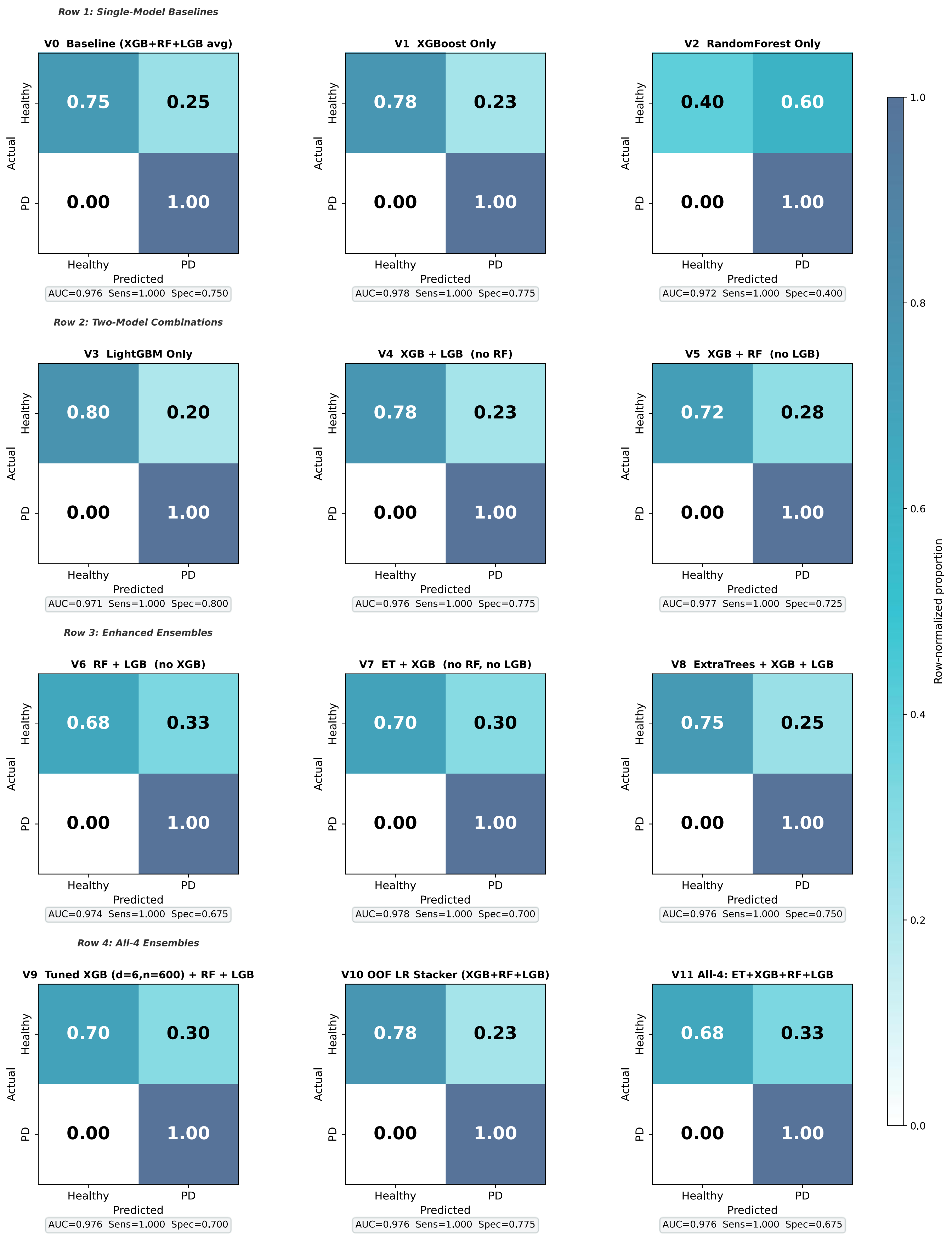

**Figure S2:** Row-normalized confusion matrices for all predefined M2 classifier configurations evaluated at the PATNO level (4×3 grid). Variants included single-model baselines (XGBoost, Random Forest, LightGBM), two-model ensemble combinations, and extended ensembles incorporating Extra Trees and Logistic Regression (LR) stacking. Across all configurations, sensitivity remained 1.000, yielding zero false negatives for PD classification; therefore, specificity was the principal metric distinguishing model performance. The final selected configuration was V3 (LightGBM only), which achieved the highest specificity among all tested models (0.800) while maintaining perfect sensitivity, with an AUC of 0.971. The XGBoost-only model (V1) demonstrated comparable overall performance (AUC = 0.978, specificity = 0.775), whereas Random Forest-only (V2) showed substantially lower specificity (0.400). Inclusion of Random Forest in larger ensembles generally reduced specificity, while ensemble averaging and LR stacking did not outperform the final LightGBM-only configuration. Cell values represent row-normalized proportions of Healthy and PD classifications at a decision threshold of 0.5.

**Figure S3:** Systematic comparison of 12 M3 collaborative filtering variants on the held-out test set.

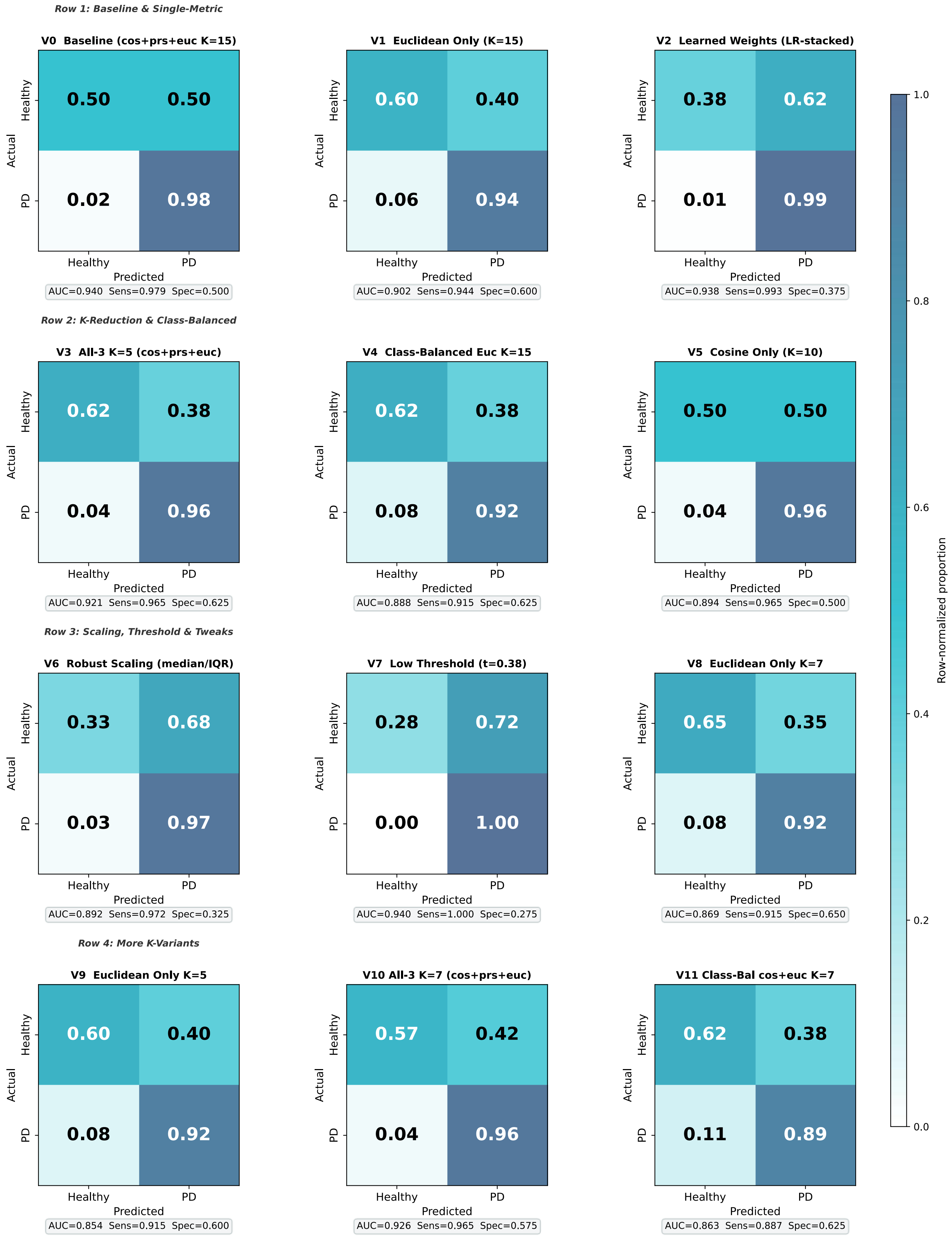

**Figure S3-** Confusion matrices for all predefined M3 configurations evaluated at the PATNO level (4×3 grid). Variants differ by similarity metric (cosine, Pearson, Euclidean), neighborhood size (K=15, 7, 5), weighting strategy (uniform, distance-weighted, learned LR-stacked weights), scaling method, and class-balancing adjustments. Across variants, sensitivity remained consistently high, whereas specificity showed substantial variability and served as the main discriminator of performance. The baseline configuration (V0; K=15, combined metrics) achieved high sensitivity (0.979) but limited specificity (0.500). Reducing neighborhood size to K=5 and averaging cosine, Pearson, and Euclidean similarities (V3) improved specificity to 0.625 while maintaining strong sensitivity (0.965) and balanced accuracy (0.795), leading to its selection as the final M3 configuration. Learned weighting and threshold modifications increased sensitivity but further reduced specificity, limiting practical utility. Cell values represent counts of Healthy and PD classifications at a 0.5 decision threshold.

**Figure S4:** ROC comparison of top-level models and final stacked ensemble on the held-out test set.

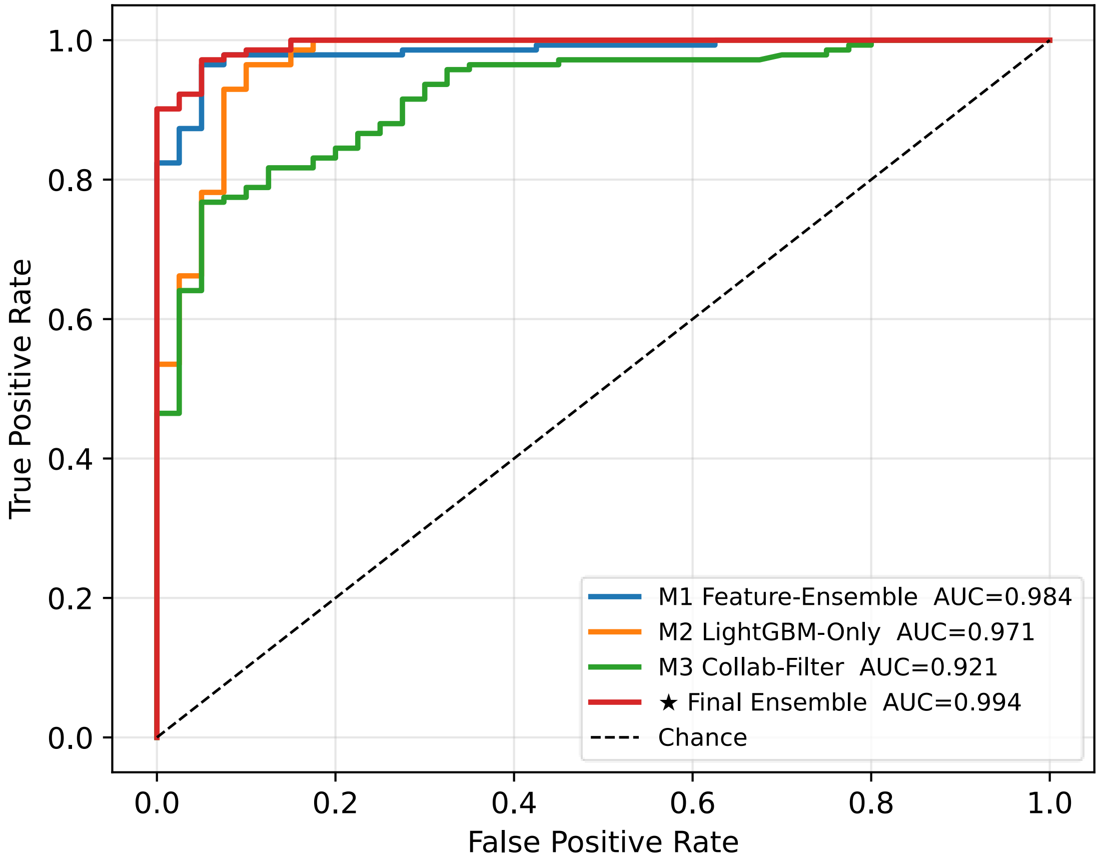


**Figure S4-** Receiver operating characteristic (ROC) curves at the PATNO level comparing M1 (feature-level ensemble), M2 (LightGBM-only), M3 (collaborative filtering), and the final stacked ensemble integrating all three sub-models. The final stacked ensemble achieved the highest discriminative performance (AUC = 0.994), followed by M1 (AUC = 0.984), M2 (AUC = 0.971), and M3 (AUC = 0.921). Although all models demonstrated strong classification capability, the stacked ensemble consistently outperformed the individual sub-models by leveraging complementary predictive information across approaches. The dashed diagonal line represents chance performance (AUC = 0.50). These findings highlight the effectiveness of the meta-stacking framework in improving overall robustness and classification accuracy beyond any single component model.

**Figure S5:** Example of a downloadable report showing a low-risk (healthy/no PD) prediction generated by the web application.

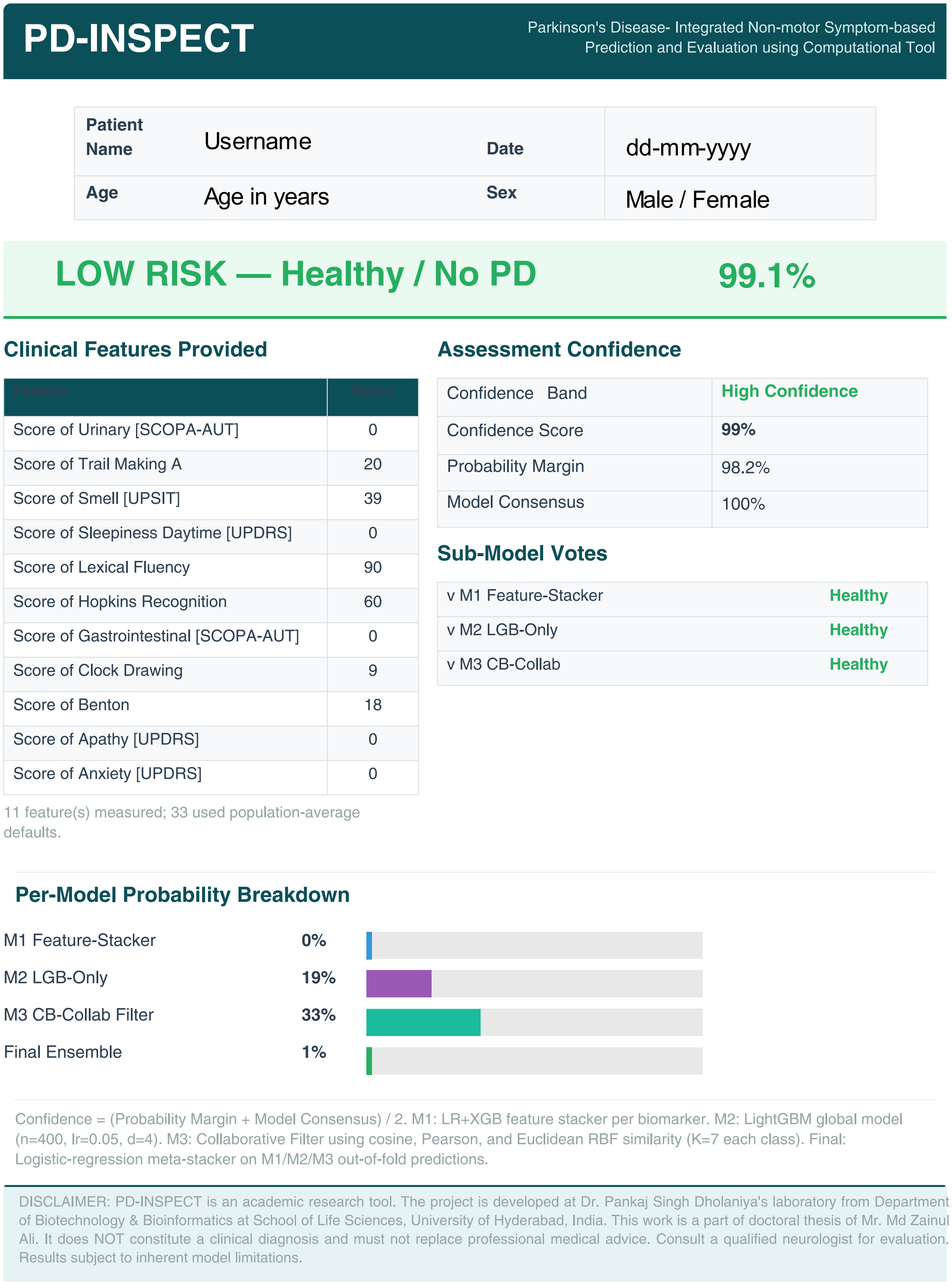

### PD-INSPECT

Parkinson's Disease- Integrated Non-motor Symptom-based Prediction and Evaluation using Computational Tool

| | | | |
|---|---|---|---|
| **Patient Name** | Username | **Date** | dd-mm-yyyy |
| **Age** | Age in years | **Sex** | Male / Female |

**LOW RISK — Healthy / No PD** **99.1%**

#### Clinical Features Provided

| Feature | Score |
|---|---|
| Score of Urinary [SCOPA-AUT] | 0 |
| Score of Trail Making A | 20 |
| Score of Smell [UPSIT] | 39 |
| Score of Sleepiness Daytime [UPDRS] | 0 |
| Score of Lexical Fluency | 90 |
| Score of Hopkins Recognition | 60 |
| Score of Gastrointestinal [SCOPA-AUT] | 0 |
| Score of Clock Drawing | 9 |
| Score of Benton | 18 |
| Score of Apathy [UPDRS] | 0 |
| Score of Anxiety [UPDRS] | 0 |

11 feature(s) measured; 33 used population-average defaults.

#### Assessment Confidence

| | |
|---|---|
| Confidence Band | **High Confidence** |
| Confidence Score | **99%** |
| Probability Margin | 98.2% |
| Model Consensus | 100% |

#### Sub-Model Votes

| | |
|---|---|
| v M1 Feature-Stacker | **Healthy** |
| v M2 LGB-Only | **Healthy** |
| v M3 CB-Collab | **Healthy** |

#### Per-Model Probability Breakdown

| | |
|---|---|
| M1 Feature-Stacker | **0%** |
| M2 LGB-Only | **19%** |
| M3 CB-Collab Filter | **33%** |
| Final Ensemble | **1%** |

Confidence = (Probability Margin + Model Consensus) / 2. M1: LR+XGB feature stacker per biomarker. M2: LightGBM global model (n=400, lr=0.05, d=4). M3: Collaborative Filter using cosine, Pearson, and Euclidean RBF similarity (K=7 each class). Final: Logistic-regression meta-stacker on M1/M2/M3 out-of-fold predictions.

DISCLAIMER: PD-INSPECT is an academic research tool. The project is developed at Dr. Pankaj Singh Dholaniya's laboratory from Department of Biotechnology & Bioinformatics at School of Life Sciences, University of Hyderabad, India. This work is a part of doctoral thesis of Mr. Md Zainul Ali. It does NOT constitute a clinical diagnosis and must not replace professional medical advice. Consult a qualified neurologist for evaluation. Results subject to inherent model limitations.

**Figure S5-** Example of a downloadable report generated by the web application for a dummy input case classified as low risk (healthy / no Parkinson's disease). The report summarizes the user-provided clinical features, overall risk interpretation, confidence metrics, and sub-model outputs. The displayed name, age, and sex are fictional placeholder fields included only to illustrate the report layout and are not derived from any real individual. User-entered information is not permanently stored by the application and remains under the control of the user. This figure is included solely to demonstrate the interface-generated output; the displayed values are representative and may vary according to the features entered by the user.
